# The association between adiposity and atypical energy-related symptoms of depression: a role for metabolic dysregulations

**DOI:** 10.1101/2022.08.16.22278833

**Authors:** Tahani Alshehri, Dennis O Mook-Kanamori, Renée de Mutsert, Brenda WJH Penninx, Frits R Rosendaal, Saskia le Cessie, Yuri Milaneschi

## Abstract

**Background:** Adiposity has been shown to be linked with atypical energy-related symptoms (AES) of depression. We used genomics to separate the effect of adiposity from that of metabolic dysregulations to examine whether the link between obesity and AES is dependent on the presence of metabolic dysregulations.

**Method:** Data were from NEO (*n*=5734 individuals) and NESDA (*n*=2238 individuals) cohorts, in which the Inventory of Depressive Symptomatology (IDS-SR30) was assessed. AES profile was based on four symptoms: increased appetite, increased weight, low energy level, and leaden paralysis. We estimated associations between AES and two genetic risk scores (GRS) indexing increasing total body fat with (metabolically unhealthy adiposity, GRS-MUA) and without (metabolically healthy adiposity, GRS-MHA) metabolic dysregulations.

**Results:** GRS-MUA and GRS-MHA were both associated with higher total body fat in NEO study, but divergently associated with biomarkers of metabolic health (e.g. fasting glucose and HDL) in both cohorts. In the pooled results, per standard deviation, GRS-MUA was specifically associated with a higher AES score (β=0.03, 95%CI: 0.01; 0.05), while there was no association between GRS-MHA and AES (β=-0.01, 95%CI: -0.03; 0.01).

**Conclusion:** These results suggest that the established link between adiposity and AES profile emerges in the presence of metabolic dysregulations, which may represent the connecting substrate between the two conditions.

## Introduction

The bidirectional relationship between obesity and depression has been well-established [1]: the presence of one of these conditions increases the risk of developing the other [2-5]. There is some evidence for a causal role of obesity in developing depression, though much still has to be elucidated [6, 7]; not every individual with depression is obese, and not every obese individual is depressed. The association between obesity and depression is complicated by heterogeneity on both sides.

Obesity is a metabolically complex and heterogenous condition. One type of obesity, known as “metabolically unhealthy”, is interwoven with cardiometabolic diseases, endocrinological alteration, and inflammation [8]. However, about 30 % of obese individuals are “metabolically healthy” [9], and excess total body fat is disconnected from these metabolic alterations [8]. A previous study by Ji et al., which combined data from genome-wide association studies on total body fat percentage and biomarkers of metabolic health, identified 14 single nucleotide polymorphisms (SNPs) associated with increased total body fat and a favourable metabolic profile characterised by higher circulating levels of HDL-cholesterol, and lower levels of triglycerides [10].

Similar to obesity, depression is a heterogeneous disorder. Individuals with a diagnosis of depression may express different symptom profiles that, in turn, are linked to different metabolic adversities. Emerging evidence [1, 11] indicates that the overlap between obesity and depression is stronger in individuals expressing atypical depressive symptoms related to altered energy intake/output balance, such as increased sleepiness, increased appetite, increased weight, low energy level and leaden paralysis. Consistently, in our earlier work [12], the four most strongly associated symptoms with increased total body fat were atypical energy-related symptoms (AES), namely increased appetite, leaden paralysis, low energy level, and increased weight. This connection is also supported by large-scale genomics studies showing genetic covariance between metabolic traits and these AES [13, 14].

The mechanism underlying the relationship between obesity and specific depressive symptoms known as atypical energy-related symptoms (AES) profile is unknown. We expect that metabolic dysregulations may represent the shared link connecting obesity with the AES profile. Studies have shown that the atypical energy-related symptom profile is associated with an adverse immuno-metabolic profile, such as BMI and CRP [15, 16], and biomarkers of neurotoxicity (kynurenine and quinolinic acid) related to low grade inflammation [17]. In the present study, we used genomics to separate the effect of adiposity from that of metabolic dysregulations to examine whether the link between obesity and AES is dependent on metabolic dysregulations. We used the same genetic instruments applied by Tyrrell et al. [7] to inspect the causal role of adiposity in the development of depression in the UK Biobank. They used two genetic risk scores (GRS, reflecting an individual’s genetic liability for a given trait) with a similar effect on total body fat but an opposing relationship with metabolic dysregulations (one predicting high total body fat without metabolic dysregulations and the other predicting high total body fat with metabolic dysregulations). The authors could not observe different patterns of associations between the two GRS and overall depression [7] but were unable to analyse specific depression symptom profiles. We expect that the association may differ when focusing on specific depressive symptom profiles.

For the current study, we used two large datasets from The Netherlands Epidemiology of Obesity study (NEO study, a population-based cohort including >6600 participants with oversampling of overweight and obese individuals) and from the Netherlands Study of Depression and Anxiety (NESDA, a prospective cohort enriched with ∼3000 participants with depressive disorders). In these studies, we derived two GRS: 1) a GRS of metabolically healthy adiposity (GRS-MHA), consisting of the SNPs associated with higher total body fat but a favourable metabolic profile identified by Ji et al. [10]; (2) a GRS of metabolically unhealthy adiposity (GRS-MUA), linked to higher adiposity and unfavourable metabolic profile based on a GWAS of BMI (See method section and appendix 1) [7, 10, 18]. We hypothesised that two GRS scores, built to index consistent association with total body fat but opposite direction associations with biomarkers of metabolic health (e.g., HDL-cholesterol and fasting glucose), and AES (i.e., increased appetite, increased weight, low energy level, and leaden paralysis). In particular, we expected that AES profile to be specifically linked with GRS-MUA reflecting increased adiposity accompanied by metabolic dysregulations.

## Method

### Study cohorts

#### The Netherlands Epidemiology of Obesity (NEO) study

NEO study is a population-based cohort study including 6671 men and women aged 45 to 65 years [19]. All inhabitants with a self-reported body mass index (BMI) of 27 kg/m^2^ or higher and living in the greater area of Leiden, the Netherlands, were eligible to participate in the NEO study. In addition, all inhabitants aged between 45 and 65 years from one adjacent municipality (Leiderdorp, the Netherlands) were invited to participate irrespective of their BMI, allowing for a reference distribution of BMI. Prior to the study visit, participants completed questionnaires at home with respect to demographic, lifestyle, and clinical information. Participants visited the NEO study centre after an overnight fast for an extensive physical examination, including anthropometry. The NEO study was approved by the medical ethics committee of Leiden University Medical Center (LUMC) and all participants gave written informed consent. This analysis included 5734 unrelated participants of European ancestry with available genetic and phenotypic information.

#### Netherlands Study of Depression and Anxiety (NESDA)

NESDA is an ongoing longitudinal cohort study that aims to describe the long-term course and consequences of depression and to examine its interaction with biological and psychosocial factors [20]. At baseline, 2981 individuals aged 18 through 65 years with depressive and/or anxiety disorders (confirmed by the Composite International Diagnostic Interview (CIDI, version 2.1.)) and healthy controls were included from the community, primary care, and secondary care settings between 2004 and 2007. The assessment included a diagnostic interview to assess the presence of depressive and anxiety disorders, a medical exam, and several questionnaires on symptom severity, other clinical characteristics and lifestyle. Participants were followed-up during four biannual assessments. For the current study, we used data from unrelated individuals of European ancestry with genetic information at the baseline data (*n*=2238) and 4 subsequent follow-up waves in which IDS-SR30 symptoms were assessed (total observations=11152). The research protocol of NESDA was approved by the medical ethical committees of the following participating universities: Leiden University Medical Centre, Vrije University Medical Centre, and University Medical Centre Groningen.

### Genetic risk scores

Genotyping, quality control, and imputation of GWAS data for both cohorts were previously described in detail [21, 22] (Appendix 2). In each cohort, we created two genetics risk scores (GRS) following the procedure previously proposed by Tyrrell et al. [7] (Appendix 1): the first one is metabolically healthy adiposity (GRS-MHA) included the 14 SNPs that were identified by Ji et al. and associated with higher total body fat but with a favourable metabolic profile (Appendix 1) [10]. The second GRS (the metabolically unhealthy adiposity (GRS-MUA)) included 76 SNPs that were linked to higher adiposity and unfavourable metabolic profile GRS index an individual’s lifetime genetic liability for a certain trait and are built as weighted sums of genetic variants associated with that trait. For each individual, the number of trait-increasing alleles carried at each SNP (0,1 or 2) is weighted for the effect size of that SNP in a GWAS of the trait of interest and then summed. In each cohort, the two GRS were standardized to a mean of zero and a standard deviation of one.

### Atypical energy-related depressive symptoms (AES)

As described in a previous study [15], the AES profile was based on the sum score of items extracted from the Inventory of Depressive Symptomatology (IDS-SR30)). The IDS-SR30 assesses (via a 4-points likert scale) the presence of 30 depressive symptoms during the last week and their severity [23]. The symptoms used in the AES included the first four top-ranking symptoms associated with total body fat in a previous analysis in the NEO study [12], namely increased appetite, leaden paralysis, low energy level, and increased weight. Increased sleepiness, previously included among atypical energy-related symptoms [15], was not among the top-ranking body-fat related symptoms and was not considered in primary analyses. In NESDA, we used baseline and four follow-up waves. AES scores at each wave were averaged in order to index the participant’s long-term exposure to depressive symptoms. In each cohort, the AES score was standardized to a mean of zero and a standard deviation of one.

### Total body fat and biomarkers of metabolic health

To benchmark the relationship between the two GRS and the total body fat and blood biomarkers of metabolic health, we used measurements of total body fat (i.e., total body fat was only available in the NEO study), triglyceride, LDL-cholesterol, HDL-cholesterol (i.e., lipid profile), and fasting glucose (i.e., glucose profile). Measurements details about biomarker of metabolic health is provided in Appendix 3.

### Statistical analysis

A schematic representation of the main elements of the study structure and the two analytical steps is depicted in Figure 1.

**Figure 1.**
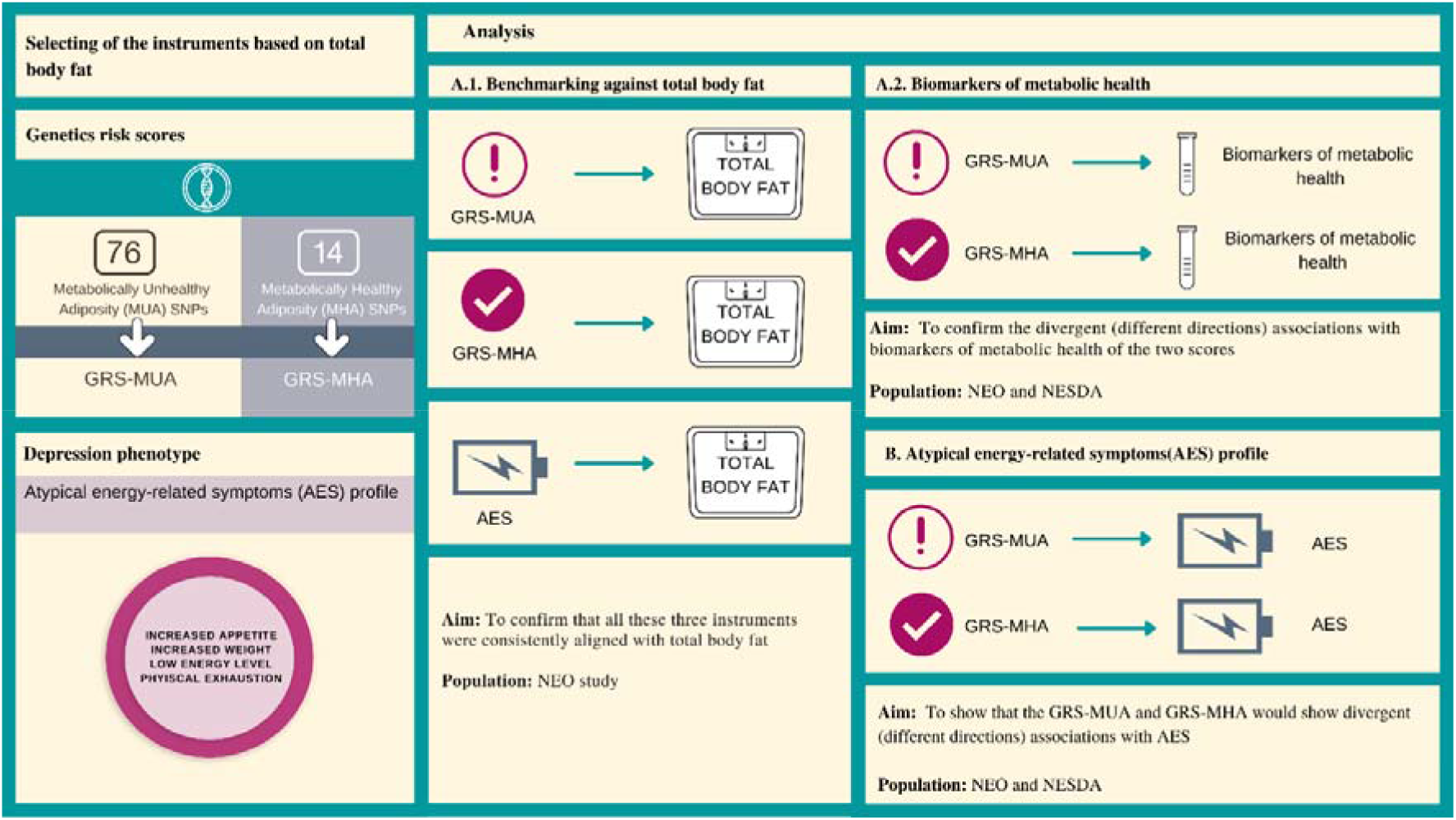
A schematic representation of the main elements of the study structure and the two analytical steps GRS-MUA: genetic risk score-metabolically unhealthy adiposity. GRS-MHA: genetic risk score: metabolically healthy adiposity. AES: atypical energy-related depressive symptoms. NEO study: The Netherlands epidemiology of obesity study. NESDA: The Netherlands study of depression and anxiety.

#### A. Benchmarking of GRS-MUA, GRS-MHA and AES against total body fat and biomarkers of metabolic health

This step consists of two parts (A.1 and A.2) (Figure 1). In the first part of step one (A.1), the associations of GRS-MUA, GRS-MHA and AES with total body fat were investigated in the NEO study. This step aimed to validate that the increase in all three instruments were associated with higher total body fat as the benchmark measure for adiposity. In the second part of step one (A.2), we estimated the association of GRS-MUA and GRS-MHA with the following biomarkers of metabolic health: triglyceride, LDL-cholesterol, HDL-cholesterol, and fasting glucose both in NEO and NESDA cohorts. The aim was to validate the different directions associations with biomarkers of metabolic health of the two GRS (GRS-MUA and GRS-MHA). Associations were estimated with linear regression models adjusted for age, sex and genetic ancestry-informative principal components. A.1 analyses were run only in NEO (due to availability of body fat measure); A.2 analyses were run in parallel in NEO and NESDA and study-specific estimates were pooled using a fixed-effect meta-analysis.

#### B. Association between GRS-MUA, GRS-MHA and Atypical energy-related symptom profile (AES)

In this main step, we estimated the association of GRS-MUA and GRS-MHA with AES. The aim of these analyses was to show divergent associations, consistently with the associations with metabolic biomarkers in A.2. GRS-MUA would be expected to show a positive association with AES, and GRS-MHA would be expected to show a negative association with AES. As in A.2, we used linear regression models adjusted for age, sex and genetic ancestry-informative principal components, and we pooled estimates obtained in NEO and NESDA using fixed-effect meta-analysis. Furthermore, we added two sensitivity analyses in which we first investigated the impact of the inclusion of increased sleepiness symptom among atypical energy-related symptom profile (i.e., by adding it as an extra symptom to the score) on the results. Second, to further confirm the specificity of the associations detected for AES, we derived similarly to previous work [15-17] a melancholic symptom profile score (0-24 range) including the following melancholic features [24]: diurnal variation (mood worse in the morning), early morning awakening, distinct quality of mood, excessive guilt, decreased appetite, decreased weight, psychomotor agitation and psychomotor retardation. All analyses were done using R version 4.0.2, and for the meta-analysis step, package (rmeta) was used.

## Results

The baseline characteristics for 5734 participants of the NEO study and 2238 participants of the NESDA included in this study are shown in Supplemental Table 1. The median of the AES was 1 point (25th-75th percentiles: 0-3), while the median of AES was 2 points (25th-75th percentiles: 1-3.6).

### A. Benchmarking against total body fat and biomarkers of metabolic health

The analyses in the first part (A.1) were done only in the NEO study. All three instruments (GRS-MUH, GRS-MHA, and AES) were associated with increased total body fat in the same direction. Effect estimate (β) in percentage total body fat per standard deviation (SD) increase of 1) GRS-MUA equal to: 0.23% (95% CI: 0.08; 0.39), 2) GRS-MHA 0.31% (95% CI: 0.15; 0.46), and 3) AES 1.43% (95% CI: 1.27; 1.58). Supplemental Table 2 shows the results of the linear regression analysis of the associations between the three instruments (GRS-MUA, GRS-MHA, and AES) and total body fat in the NEO study. Figure 2 depicts the predicted values of total body fat as a function of above mentioned three instruments. These results confirmed that the two GRS and the AES profile were consistently aligned to body fat. Then, the second part of this step (A.2) confirmed that the GRS-MUA and GRS-MHA were differently associated with the biomarkers of metabolic health in NEO and NESDA cohorts (Supplemental Table 2 for cohort specific association). Figure 3 depicts the pooled (and supplemental table 3 shows cohort-specific) effect estimates and 95% confidence intervals for the association of the two genetics risk scores and the biomarkers of metabolic health. GRS-MUH was associated with an adverse metabolic profile such as (per SD)) in higher fasting glucose 0.03 mmol/L (95% CI: 0.01; 0.05) and lower HDL-cholesterol -0.02 mmol/L (−0.04; 0.00). The GRS-MHA was linked to a favourable metabolic profile, such as (per SD) lower fasting glucose -0.03 mmol/L (−0.05; 0.00) and higher HDL cholesterol 0.07 mmol/L (0.05; 0.09).

**Figure 2.**
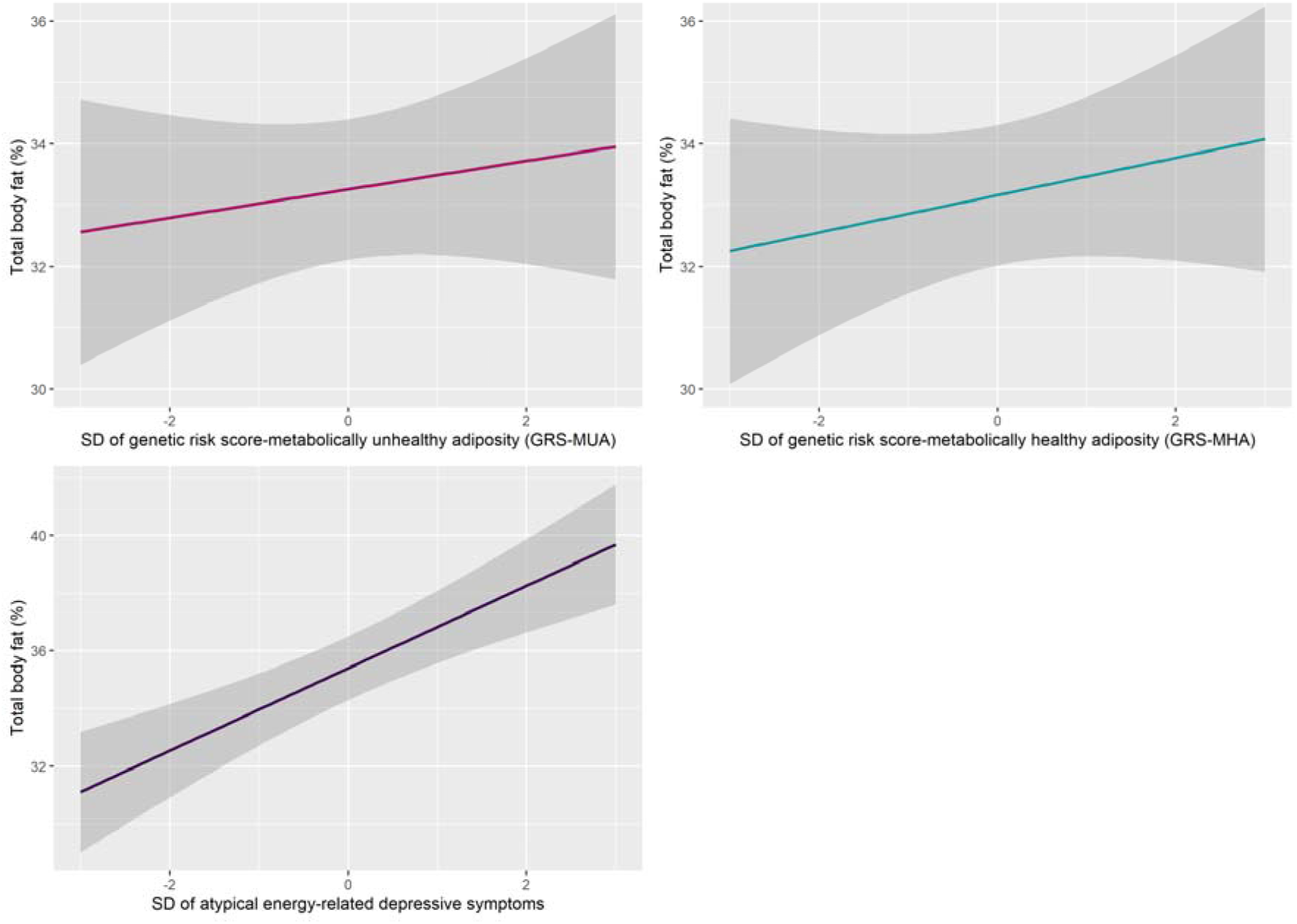
Predicted values of total body fat in the NEO study as function of the GRS-MUA, GRS-MHA, and AES SD: standard deviation. AES: Atypical energy-related symptom profile: a sum score of the four depressive symptoms, increased appetite, increased weight, low energy level, and leaden paralysis. The grey area represents 95% confidence interval.

**Figure 3.**
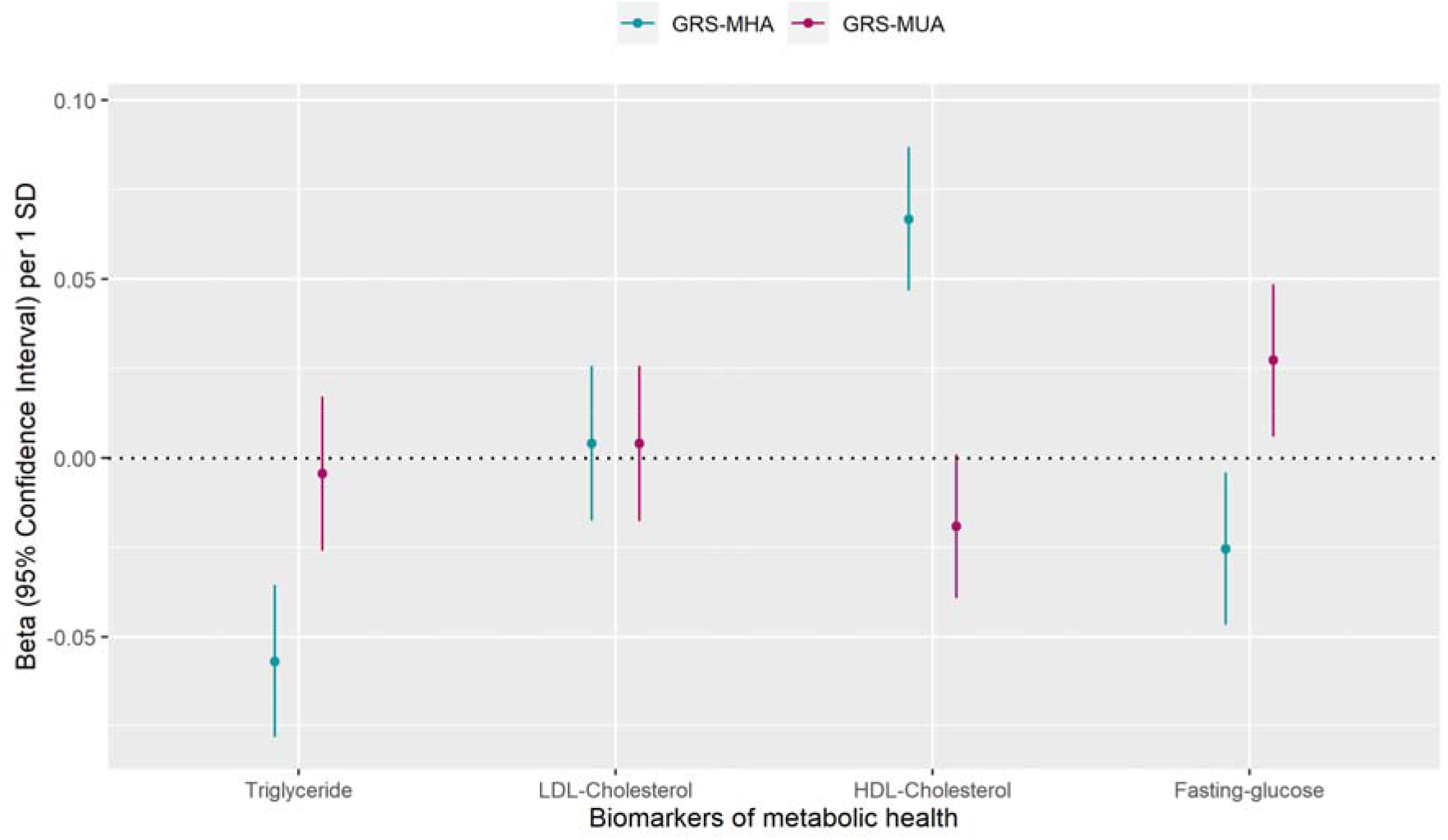
Pooled results of effect estimates of the linear regression between the genetics instruments (GRS-MUA, GRS-MHA) and biomarkers of metabolic health, model adjusted for age, sex, and genetic ancestry-informative principal components GRS-MUA: Genetics risk score metabolically unhealthy adiposity, GRS-MHA: Genetics risk score metabolically healthy adiposity. SD: standard deviation

### B. Atypical energy-related symptom profile

Finally, we examined the association between the two genetic risk scores (GRS-MUA, GRS-MHA) and the AES profile. Figure 4 shows pooled estimates and 95% CIs, and supplemental table 4a shows cohort-specific effect estimates and 95% CIs of the associations with AES from linear regression models adjusted for age, sex, and genetic ancestry-informative principal components. GRS-MUA was specifically associated with higher AES (per SD) 0.03 (95% CI: 0.01;0.05); in contrast, GRS-MHA was not associated with AES -0.01 (−0.03;0.01). Adding increased sleepiness to the AES yielded similar results indicating that a substantial proportion of genetic co-variance between GRS-MUA and AES was already captured by the four symptoms of increased appetite, increased weight, low energy level, and leaden paralysis. Figure 4 and Supplemental Table 5a show that neither GRS-MUA nor GRS-MHA were associated with melancholic symptom profile. This finding suggests that the detected link between GRS-MUA and AES is specific for this symptom profile. Finally, we repeated this step (B) using BMI-weighted analyses in the NEO study. Since NEO is a population-based study with oversampling of individuals with a BMI > 27 kg/m2, a weighted analyses were performed as sensitivity analyses. The weighting factor is based on BMI distribution in the general Dutch population to make our results generalizable to the Dutch population. This procedure did not substantially change the results (Supplemental Table 4b and 5b).

**Figure 4.**
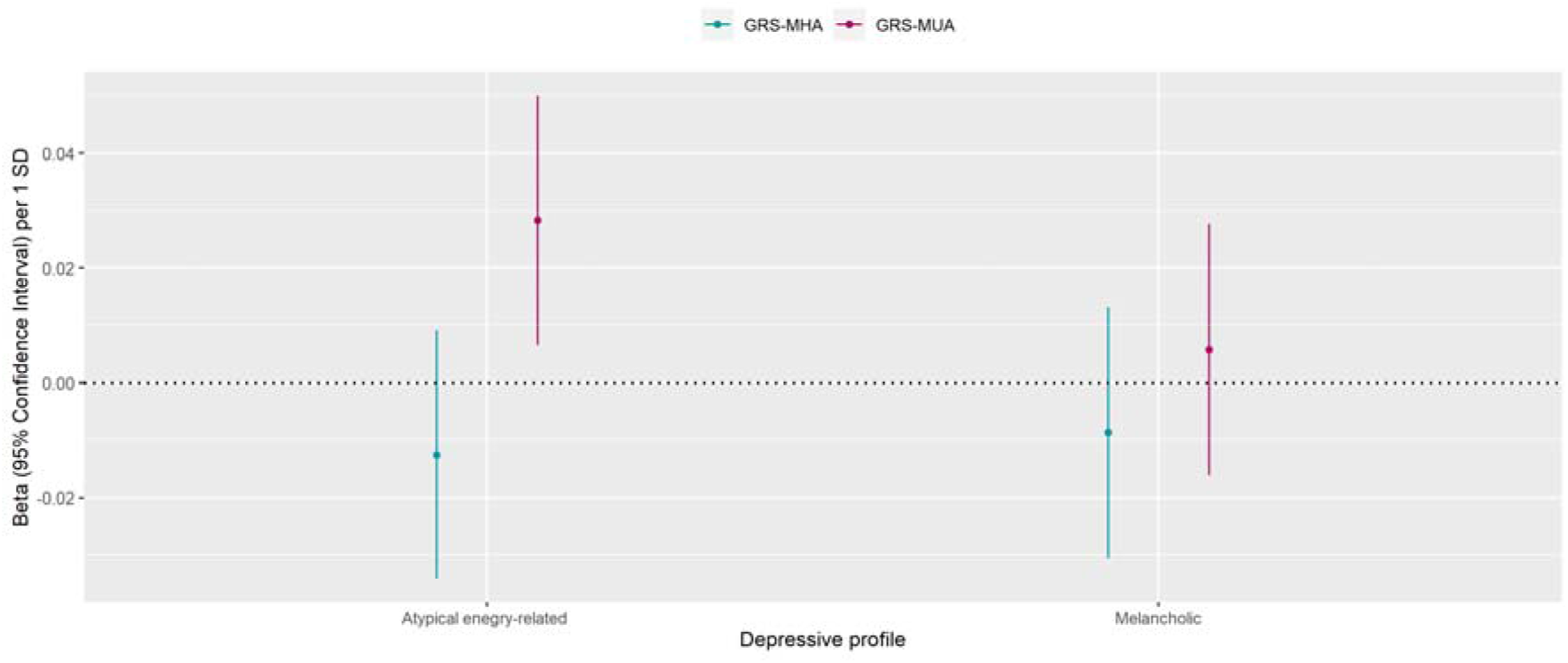
Pooled results of effect estimate of the linear regressions between the genetics instruments (GRS-MUA, GRS-MHA) and atypical energy related symptoms and melancholic symptoms profile, model adjusted for age, sex, and genetic ancestry-informative principal components. GRS-MUA: Genetics risk score metabolically unhealthy adiposity, GRS-MHA: Genetics risk score metabolically healthy adiposity. SD: standard deviation. Atypical energy-related symptom profile: a sum score of the four depressive symptoms, increased appetite, increased weight, low energy level, and leaden paralysis. Melancholic depressive symptoms profile: a sum score of the symptoms, decreased appetite, decreased weight, early morning awakening, mood variation in relation to the time of the day, distinct quality of mood, excessive guilt, psychomotor agitation, and psychomotor retardation

## Discussion

This study investigated whether the established link between adiposity and AES of depression is rooted in underlying metabolic dysregulations. For that, we uncoupled the effect of adiposity from that of metabolic dysregulations. We studied the relationships between two adiposity increasing genetic risk scores (i.e., GRS-MUA and GRS-MHA) and AES. Both genetic instruments used in this study increased the predisposition to high adiposity. The discrepancy between them is that GRS-MUA also increases the predisposition to metabolic dysregulations, and GRS-MHA associates with a favourable metabolic profile. We found that GRS-MUA and GRS-MHA both predicted a high total body fat level but divergently associated with metabolic dysregulations and AES. In particular, GRS-MUA was specifically associated with higher AES scores. Overall, these results suggest that the established link between adiposity and atypical energy-related depressive symptoms emerges in the presence of metabolic dysregulations, which may represent the connecting substrate between the two conditions.

The mechanisms underlying this relationship between adiposity and this specific depression profile are unknown. The recently introduced transdiagnostic model of immuno-metabolic depression (IMD) [25] suggests that inflammatory and metabolic dysregulations act as a shared substrate influencing the development of specific behavioural symptoms common to depression and obesity. For instance, alterations in central signalling of leptin and insulin may associate with shifting body energy balance from expenditure to accumulation, favouring the development of hyperphagia, present in both obesity and atypical form of depression. Finally, these metabolic dysregulations have been hypothesised to be the link between depression and cardiovascular diseases. For example, immuno-metabolic dysregulations commonly linked to CVD, such as triglyceride, IL-6, and CRP, were causally related to depression [26]. Interestingly, a recent study showed that higher inflammation measured by IL-6 activity is a potential causal for a specific symptom profile of depression, such as sleep problems or fatigue [27].

Other mechanisms related to body fat but not associated with immuno-metabolic biological alterations (e.g. weight shame [28], body image dissatisfaction [29]) may play a role in developing and experiencing depression. However, considering that in our results, GRS-MHA was not related to higher AES, these alternative mechanisms seem less likely. A previous individual-participants meta-analysis study [30] pooled data from 8 studies (n>30000) to test the relationship between metabolically healthy adiposity and depression. They divided individuals into four groups, non-obese metabolically healthy (reference), non-obese metabolically unhealthy, obese metabolically healthy, and obese metabolically unhealthy. They found an increased risk of depression in all three categories in comparison to the reference [30]. This might mean that the body image dissatisfaction explanation may be still valid for the other types of depression.

The present findings highlight the importance of resolving depression heterogeneity when examining its biology. Tyrrell et al. [7] and Marten et al. [31] inspected the causal role of adiposity (via two instrumental variables, metabolically unhealthy adiposity GRS and metabolically healthy adiposity GRS) in the development of depression in the UK Biobank. For example, Tyrrell et al. [7] hypothesised that the GRS-MUA would be associated with depression due to the underlying metabolic dysregulation and GRS-MHA would not be associated with depression for the link with the favourable metabolic profile. Instead, they found that both GRS-MUA and GRS-MHA were associated with depression. The results of [7] and [31] exemplify how depression heterogeneity hinders efforts to identify its biological underpinnings. In this work, we found a positive association between GRS-MHU and AES and a negative association between GRS-MHA and AES, which was in the direction initially hypothesised by Tyrrell et al. by focusing on a specific depressive symptom profile. The present findings are consistent with previous genetic studies that showed the AES was associated with the genetic risk scores that related to a higher risk of adiposity and its related immunometabolic dysregulations such as GRS of BMI and CRP [32]. Moreover, two large scale genetics studies in > 30000 individuals from the UK Biobank [33] and >26000 individuals from Psychiatric Genomics Consortium [13] found a genetics overlap between adiposity related traits such as BMI, leptin and CRP levels and AES (e.g., increased weight). Overall, evidence from those previous studies and the present one support the hypothesis that the link between adiposity and AES is driven by immune-metabolic dysregulation [25].

The strengths of the present study are, first, we used a large sample size (n> 7000) by combining participants from two cohorts. Second, both the NEO study (i.e., a population-based study that focuses on obesity) and the NESDA cohort (i.e., a clinical cohort study that focuses on depression) have similar genetics and symptoms instruments and detailed biomarkers of metabolic health. However, some limitations need to be addressed. First, based on the different sample sizes between NEO and NESDA, the meta-analysed results of the pooled analyses are driven by the largest study. Nonetheless, the results in both studies were similar. Second, considering the observational design of the study, causality questions about the association between the two genetics risk scores and AES cannot be answered in this study. Third, GRS were derived using summary genotype data and GWAS summary statistics obtained from subjects of European ancestry GWASs, which make our results not fully generalizable to other ethnicities.

This study showed that the established link between adiposity and atypical energy-related depressive symptoms emerges in the presence of metabolic dysregulation. This supports the hypothesis that metabolic dysregulation represents a key connecting mechanism between adiposity and a specific form of depression. Albeit health care providers shift from assessing adiposity based on BMI solely by incorporating waist circumference and lipid profile to diagnose the overall health profile, less has been done regarding the depression heterogeneity. Monitoring the metabolic health of patients who express atypical energy-related symptomatology could be beneficial to prevent the development of cardiometabolic disorders.

## Data Availability

All data produced in the present study are available upon reasonable request to the authors

## NESDA

The infrastructure for the NESDA study (www.nesda.nl) is funded through the Geestkracht program of the Netherlands Organization for Health Research and Development (ZonMw, grant number: 10-000-1002) and financial contributions by participating universities and mental health care organizations (VU University Medical Center, GGZ inGeest, Leiden University Medical Center, Leiden University, GGZ Rivierduinen, University Medical Center Groningen, University of Groningen, Lentis, GGZ Friesland, GGZ Drenthe, Rob Giel Onderzoekscentrum).

## NEO study

The NEO study is supported by the participating Departments, Division, and Board of Directors of the Leiden University Medical Center, and by the Leiden University, Research Profile Area Vascular and Regenerative Medicine. DOM-K is supported by Dutch Science Organization (ZonMW-VENI Grant No. 916.14.023). We express our gratitude to all participants of the Netherlands Epidemiology of Obesity (NEO) study, in addition to all participating general practitioners. We furthermore thank P.R. van Beelen and all research nurses for collecting the data, P.J. Noordijk and her team for sample handling and storage, and I. de Jonge for data management of the NEO study. Finally, we would like to express our gratitude to Dr R. Noordam for extracting the genetics data in this study and answering our related questions.

