## Supplemental File # 1 for "The association between adiposity and atypical energy-related symptoms of depression: a role for metabolic dysregulations"

**Table of Contents**

**Appendix 1**. Genetic risk scores	2

**Appendix 2.** Genetic data technical report (genotyping and imputation)	3

Genotyping and Imputation in NEO study	3

Genotyping and Imputation in NESDA	3

**Appendix 3.** Total body fat and biomarkers of metabolic health	4

**Supplemental Table 1.** Characteristics of the study population NESDA and NEO cohorts	6

**Supplemental Table 2.** Results of the linear regression analysis of the association between the three instruments GRS-MUA, GRS-MHA, AES and body fat in NEO study.	8

**Supplemental Table 3.** Results of the linear regression analysis of the association between the genetics instruments (GRS-MUA, GRS-MHA) and biomarkers of metabolic health.	9

**Supplemental Table 4a**. Results of the linear regression analysis of the association between the genetics instruments (GRS-MUA, GRS-MHA) and atypical energy-related depressive symptoms.	12

**Supplemental Table 4b.** Results of the linear regression analysis of the association between the genetics instruments (GRS-MUA, GRS-MHA) and atypical energy-related depressive symptoms (weighted analysis).	13

**Supplemental Table 5a.** Results of the linear regression analysis of the association between the genetics instruments (GRS-MUA, GRS-MHA) and melancholic depressive symptoms.	14

**Supplemental Table 5b.** Results of the linear regression analysis of the association between the genetics instruments (GRS-MUA, GRS-MHA) and melancholic depressive symptoms.	15

**References**	16

**Appendix 1**. Genetic risk scores

In each cohort (i.e., NESDA and NEO), we created two genetics risk scores (GRS): the first one is metabolically healthy adiposity (GRS-MHA) included the 14 SNPs [1] that associated with higher total body fat but a favourable metabolic profile. Ji et al [1] identified these 14 SNPs in three steps analyses. First, SNPs related to increase total body fat were identified based on a GWAS of total body fat in more than 442,000 individuals in the UK Biobank. Second, multivariate GWAS of metabolic biomarkers performed based on the summary statistics of the GWASs of the following metabolic biomarkers: total body fat (n=120000) [2], HDL-cholesterol (n=99900) [3], adiponectin (n = 29,400) [4], sex hormone-binding globulin (n=21800) [5], triglyceride (n=96600) [3], fasting insulin (n=51800) [6] and alanine transaminase (n=55500) [7]. Third, genetics variants associated with step 1 and step 2 were selected (SNPs related to metabolically healthy adiposity). The second GRS was linked to higher adiposity and unfavourable metabolic profile (metabolically unhealthy adiposity (GRS-MUA)) based on a GWAS of BMI in 339,224 individuals [8, 9], where 76 SNPs associated with metabolically unhealthy adiposity were identified. Following the procedure previously proposed by Tyrrell et al [9], we calculated GRS-MUA based on 76 SNPs (i.e., 75 SNPs were available in NEO and 72 SNPs in NESDA) [8, 9]. GRS were calculated as follows: each individual variants were recoded as 0, 1 and 2, according to the number of adiposity increasing alleles. Each variant was weighted by its effect size (β-coefficient) obtained from the primary GWAS [8], then a sum of the weighted variants was derived as previously done by Ji et al and Tyrrell et al [1, 9]. In each cohort, The two GRS were standardized to a mean of zero and a standard deviation of one, allowing interpretability.

**Appendix 2.** Genetic data technical report (genotyping and imputation)

Genotyping, quality control, and imputation of GWAS data for NEO and NESDA cohorts were previously described in detail [10, 11].

### Genotyping and Imputation in NEO study

DNA was extracted from venous blood samples obtained from the antecubital vein. Genotyping was performed in Centre National de Génotypage (Evry Cedex, France), using the Illumina HumanCoreExome-24 BeadChip (Illumina, San Diego, CA). The detailed quality-control process has previously been described [10]. Genotypes were further imputed to the 1000 Genome Project reference panel (version 3, 2011) [12] using IMPUTE (version 2.2) software [13]. No genetic variants with an imputation quality <0.4 or a minor allele frequency (MAF) <0.01 were considered for the analyses in the current study.

### Genotyping and Imputation in NESDA

Methods for biological sample collection and DNA extraction have been described previously [14]. Quality control and imputation pipelines were also previously described [11]. Briefly, 95% of the samples were genotyped on the Affymetrix 6.0 Human SNP array and the remaining on the Perlegen-Affymetrix 5.0 array. After platform-specific QC the missing SNP genotypes between each platform were imputed using the GONL (Genome of the Netherlands) [15-17] reference panel and then merged, followed by additional more stringent QC. This cross-platform GONL imputed dataset was used to identify ancestry outliers, defined based on Principal Components Analysis (PCA) by projecting 10 PCs from 1000G reference set populations on the cross-platform imputed data using the SMARTPCA program as described earlier [18, 19]. Individuals with PC values located outside of the range of European and/or British populations were defined as outliers. Upon exclusion of outliers, 10 PCs were recomputed for cross-platform imputed data to capture the variation within the Netherlands. The SNPs from the cross-platform GONL imputed dataset (~1.3M) were used for a second round of imputations to the Haplotype Reference Consortium [20] reference panel using the Michigan Imputation Server [21]. The cross-platform imputed dataset was used to build a relationship matrix measuring genetic similarity using GCTA[22], which was pruned at 0.05 threshold in order to retain unrelated participants. After application of additional post-imputation QC (MAF > 0.01, HWE-*p* > 1e-6) 87 SNPs were extracted for the present analyses (Supplemental Table 5) . All the selected SNPs had high imputation quality (<0.6).

**Appendix 3.** Total body fat and biomarkers of metabolic health

To confirm the relationship between the two GRS and the total body fat and blood biomarkers of metabolic health, we used measurements of total body fat (i.e., total body fat was only available in NEO study), and triglyceride, LDL-cholesterol, HDL-cholesterol (i.e., lipid profile), and fasting glucose (i.e., glucose profile). We additionally used HOMA of beta-cell function (HOMA-1B,) Homeostasis Model Assessment for Insulin Resistance (HOMA-IR), and HbA1c (%) that were only available in the NEO study. Total body fat was measured by Tanita bioelectrical impedance balance (TBF-310, Tanita International Division, UK). Lipid and glucose profile were measured from fasting plasma samples by using standard clinical laboratory techniques [23, 24]. From fasting glucose and insulin concentrations, we calculated the HOMA-IR and HOMA-1B as markers of hepatic insulin resistance and steady-state insulin secretion [25]. HOMA-IR was calculated as fasting insulin (µU/mL) x fasting glucose (mmol/L)/22.5 and HOMA-1B% as 20 x fasting glucose (mmol/l)-3.5 [25, 26]. In each cohort, all biomarkers of metabolic health were standardized to a mean of zero and a standard deviation for each variable of interest of one, allowing interpretability.

**Supplemental Table 1.** Characteristics of the study population NESDA and NEO cohorts

|  | NESDA | NEO |
| --- | --- | --- |
| N | 2238 | 5734 |
| Women, n (%) | 1481 (66.2) | 2980 (52.0) |
| Age (years) (mean, sd) | 42.75 (12.9) | 55.97 (5.9) |
| BMI (kg/m^2^) (mean, sd) | 25.70 (5.0) | 29.98 (4.8) |
| Waist circumference (cm) | 89.63 (14.1) | 101.96 (13.2) |
| Total body fat (%) | - | 36.3 (28.4, 43.2) |
| Glucose (mmol/L) | 5.0 (4.7,5.5) | 5.72 (1.1) |
| Serum concentration of LDL (mmol/L) | 3.16 (1.0) | 3.57 (1.0) |
| Serum concentration of HDL (mmol/L) | 1.62 (0.4) | 1.43 (0.4) |
| Serum concentration of triglyceride (mmol/L) | 1.1(0.8-1.6) | 1.25 (0.88, 1.77) |
| Atypical energy-related symptoms profile (AES) | 2 (1,3.6)  2.4 (1.92) | 1 (0, 3)  2 (1.9) |
| Melancholic symptoms profile | 3 (1.2,5.4)  3.7 (3.03) | 1 (0, 3)  2 (2.4) |

Normally distributed data are shown as mean and standard deviation (SD). Skewed distributed data are shown as median (25th, 75th percentiles). Categorical data are shown as a number and percentage (n (%)). Atypical energy-related symptom profile: a sum score of the four depressive symptoms, increased appetite, increased weight, low energy level, and leaden paralysis. Melancholic depressive symptoms profile: a sum score of the symptoms, decreased appetite, decreased weight, early morning awakening, mood variation in relation to the time of the day, distinct quality of mood, excessive guilt, psychomotor agitation, psychomotor retardation. BMI: body mass index. NESDA: Netherlands Study of Depression and Anxiety. NEO: Netherlands Epidemiology of Obesity study. GRS-MUA: Genetics risk score metabolically unhealthy adiposity, GRS-MHA: Genetics risk score metabolically healthy adiposity.

**Supplemental Table 2.** Results of the linear regression analysis of the association between the three instruments GRS-MUA, GRS-MHA, AES and body fat in NEO study.

|  | Total body fat (%)  β(95% CI) |
| --- | --- |
| **GRS-MUA** | 0.23 (0.08;0.39) |
| **GRS-MHA** | 0.31 (0.15;0.46) |
| **AES** | 1.43 (1.27;1.58) |

GRS-MUA: GRS-metabolically unhealthy adiposity. GRS-MHA:GRS-metabolically healthy adiposity. AES: Atypical energy-related symptom profile (a sum score of the four symptoms, increased appetite, increased weight, low energy level, leaden paralysis.

**Supplemental Table 3.** Results of the linear regression analysis of the association between the genetics instruments (GRS-MUA, GRS-MHA) and biomarkers of metabolic health.

|  |  | **GRS-MUA** |  | **GRS-MHA** |  |
| --- | --- | --- | --- | --- | --- |
|  |  | β(95% CI) | P-value | β(95% CI) | P-value |
| **Fasting glucose (mmol/L)** | **NEO** | 0.01 (-0.01;0.04) | 2.50 X 10^-01^ | -0.03 (-0.06;-0.01) | 1.75 X 10^-02^ |
|  | **NESDA** | 0.06 (0.02;0.10) | 4.48 X 10^-03^ | -0.01 (-0.05;0.03) | 5.23 X 10^-01^ |
|  | **Pooled** | 0.03 (0.01;0.05) | 1.17 X 10^-02^ | -0.03 (-0.05;0.00) | 1.93 X 10^-02^ |
| **HOMA-1B** | **NEO** | -0.01 (-0.04;0.01) | 3.58 X 10^-01^ | -0.04 (-0.06;-0.01) | 6.90 X 10^-03^ |
|  | **NESDA** | - |  | - |  |
|  | **Pooled** | - |  | - |  |
| **HOMA-IR** | **NEO** | -0.01 (-0.03;0.02) | 6.60 X 10^-01^ | -0.04 (-0.07;-0.01) | 1.98 X 10^-03^ |
|  | **NESDA** | - |  | - |  |
|  | **Pooled** | - |  | - |  |
| **HbA1c (%)** | **NEO** | 0.02 (-0.01;0.04) | 2.04 X 10^-01^ | -0.03 (-0.05;0.00) | 4.76 X 10^-02^ |
|  | **NESDA** | - |  | - |  |
|  | **Pooled** | - |  | - |  |
| **HDL-cholesterol (mmol/L)** | **NEO** | -0.02 (-0.04;0.01) | 1.61 X 10^-01^ | 0.07 (0.05;0.09) | 4.21 X 10^-09^ |
|  | **NESDA** | -0.03 (-0.07;0.01) | 1.93 X 10^-01^ | 0.06 (0.02;0.10) | 3.61 X 10^-03^ |
|  | **Pooled** | -0.02 (-0.04;0.00) | 6.14 X 10^-02^ | 0.07 (0.05;0.09) | 5.88 X 10^-11^ |
| **Triglycerides (mmol/L)** | **NEO** | -0.01 (-0.04;0.01) | 3.15 X 10^-01^ | -0.06 (-0.09;-0.04) | 6.18 X 10^-07^ |
|  | **NESDA** | 0.02 (-0.02;0.06) | 4.11 X 10^-01^ | -0.04 (-0.08;0.00) | 6.26 X 10^-02^ |
|  | **Pooled** | 0.00 (-0.03;0.02) | 6.88 X 10^-01^ | -0.06 (-0.08;-0.04) | 1.91 X 10^-07^ |
| **LDL-cholesterol (mmol/L)** | **NEO** | 0.02 (-0.01;0.04) | 1.62 X 10^-01^ | 0.01 (-0.02;0.04) | 4.22 X 10^-01^ |
|  | **NESDA** | -0.03 (-0.07;0.01) | 1.44 X 10^-01^ | -0.01 (-0.05;0.03) | 5.79 X 10^-01^ |
|  | **Pooled** | 0.00 (-0.02;0.03) | 7.16 X 10^-01^ | 0.00 (-0.02;0.03) | 7.15 X 10^-01^ |

HOMA-1B: HOMA of beta-cell function. HOMA-IR: Homeostasis Model Assessment for Insulin Resistance.

**Supplemental Table 4a**. Results of the linear regression analysis of the association between the genetics instruments (GRS-MUA, GRS-MHA) and atypical energy-related depressive symptoms.

|  |  | **GRS-MUA** | | **GRS-MHA** | |
| --- | --- | --- | --- | --- | --- |
|  |  | **β(95% CI)** | **p-value** | **β(95% CI)** | **p-value** |
| **AES** | **NEO** | 0.02 (0.00;0.05) | 1.11 X 10^-01^ | -0.02 (-0.05;0.01) | 1.17 X 10^-01^ |
|  | **NESDA** | 0.05 (0.01;0.09) | 2.27 X 10^-02^ | 0.01 (-0.03;0.05) | 7.02 X 10^-01^ |
|  | **Pooled** | 0.03 (0.01;0.05) | 1.06 X 10^-02^ | -0.01 (-0.03;0.01) | 2.56 X 10^-01^ |
| **AES**  **(sensitivity)** | **NEO** | 0.02 (-0.01;0.04) | 2.39 X 10^-01^ | -0.02 (-0.04;0.01) | 2.35 X 10^-01^ |
|  | **NESDA** | 0.04 (0.00;0.09) | 3.71 X 10^-02^ | 0.00 (-0.04;0.04) | 9.61 X 10^-01^ |
|  | **Pooled** | 0.02 (0.00;0.05) | 3.59 X 10^-02^ | -0.01 (-0.03;0.01) | 3.25 X 10^-01^ |

AES :Atypical energy-related symptom profile: a sum score of the four symptoms, increased appetite, increased weight, low energy level, leaden paralysis. AES (sensitivity): a sum score of the five symptoms, increased sleepiness, increased appetite, increased weight, low energy level, leaden paralysis.

**Supplemental Table 4b.** Results of the linear regression analysis of the association between the genetics instruments (GRS-MUA, GRS-MHA) and atypical energy-related depressive symptoms (weighted analysis).

|  |  | **GRS-MUA** | | **GRS-MHA** | |
| --- | --- | --- | --- | --- | --- |
|  |  | **β(95% CI)** | **p-value** | **β(95% CI)** | **p-value** |
| **AES** | **NEO** | 0.02 (-0.02;0.05) | 3.21 X 10^-01^ | -0.03 (-0.06;0.00) | 4.2 X 10^-02^ |
|  | **NESDA** | 0.05 (0.01;0.09) | 2.27 X 10^-02^ | 0.01 (-0.03;0.05) | 7.02 X 10^-01^ |
|  | **Pooled** | 0.03 (0.00;0.05) | 2.9 X 10^-02^ | -0.02 (-0.04;0.01) | 1.5 X 10^-01^ |
| **AES**  **(sensitivity)** | **NEO** | 0.02 (-0.02;0.05) | 3.08 X 10^-01^ | -0.02 (-0.05;0.01) | 1.8 X 10^-01^ |
|  | **NESDA** | 0.04 (0.00;0.09) | 3.71 X 10^-02^ | 0.00 (-0.04;0.04) | 9.61 X 10^-01^ |
|  | **Pooled** | 0.02 (0.00;0.04) | 3.0 X 10^-02^ | -0.01 (-0.04;0.01) | 3.0 X 10^-01^ |

AES :Atypical energy-related symptom profile: a sum score of the four symptoms, increased appetite, increased weight, low energy level, leaden paralysis.. AES (sensitivity): a sum score of the five symptoms, increased sleepiness, increased appetite, increased weight, low energy level, leaden paralysis.

**Supplemental Table 5a.** Results of the linear regression analysis of the association between the genetics instruments (GRS-MUA, GRS-MHA) and melancholic depressive symptoms.

|  |  | **GRS-MUA** | | **GRS-MHA** | |
| --- | --- | --- | --- | --- | --- |
|  |  | **β(95% CI)** | **p-value** | **β(95% CI)** | **p-value** |
| **Melancholic symptoms profile**  **(sensitivity)** | **NEO** | 0.00 (-0.02;0.03) | 9.40 X 10^-01^ | -0.02 (-0.04;0.01) | 1.71 X 10^-01^ |
|  | **NESDA** | 0.02 (-0.02;0.06) | 3.95 X 10^-01^ | 0.02 (-0.03;0.06) | 4.71 X 10^-01^ |
|  | **Pooled** | 0.01 (-0.02;0.03) | 6.07 X 10^-01^ | -0.01 (-0.03;0.01) | 4.34 X 10^-01^ |

Melancholic symptoms profile (sensitivity): a sum score of the symptoms, decreased appetite, decreased weight, early morning awakening, mood variation in relation to the time of the day, distinct quality of mood, excessive guilt, psychomotor agitation, psychomotor retardation.

**Supplemental Table 5b.** Results of the linear regression analysis of the association between the genetics instruments (GRS-MUA, GRS-MHA) and melancholic depressive symptoms.

|  |  | **GRS-MUA** | | **GRS-MHA** | |
| --- | --- | --- | --- | --- | --- |
|  |  | **β(95% CI)** | **p-value** | **β(95% CI)** | **p-value** |
| **Melancholic symptoms profile**  **(sensitivity)** | **NEO** | 0.00 (-0.04;0.04) | 9.16 X 10^-01^ | -0.03 (-0.07;0.00) | 8.11 X 10^-02^ |
|  | **NESDA** | 0.02 (-0.02;0.06) | 3.95 X 10^-01^ | 0.02 (-0.03;0.06) | 4.71 X 10^-01^ |
|  | **Pooled** | 0.01 (-0.02;0.04) | 5.18 X 10^-01^ | -0.01 (-0.04;0.01) | 6.93 X 10^-02^ |

Melancholic symptoms profile (sensitivity): a sum score of the symptoms, decreased appetite, decreased weight, early morning awakening, mood variation in relation to the time of the day, distinct quality of mood, excessive guilt, psychomotor agitation, psychomotor retardation.
